# Assessing the health benefits of physical activity due to active commuting in a French energy transition scenario

**DOI:** 10.1101/2022.02.17.22271146

**Authors:** Pierre Barban, Audrey de Nazelle, Stéphane Chatelin, Philippe Quirion, Kévin Jean

## Abstract

**Background:** Energy transition scenarios are prospective outlooks describing combinations of changes in socio-economic systems that are compatible with climate targets. These changes could have important health co-benefits. We aimed to quantify the health benefits of physical activity caused by active transportation on all-cause mortality in the French negaWatt scenario over the 2021-2050 period.

**Methods:** Relying on a health impact assessment framework, we quantified the health benefits of increased walking, cycling and E-biking projected in the negaWatt scenario. The negaWatt scenario assumes increases of walking and cycling volumes of +11% and +612%, respectively, over the study period.

**Results:** As compared to a scenario with no volume increase, we quantified that the negaWatt scenario would prevent 9,797 annual premature deaths in 2045 and translate into a 3-month increase of life expectancy in the general population. These health gains would generate €34 billion of economic benefits from 2045 onwards,

**Conclusions:** Increased physical activity implied in the negaWatt transition scenario would generate substantial public health benefits, which are comparable to the gain expected by large scale health prevention interventions.

## INTRODUCTION

The health impact of climate change have recently become more prominent features of climate change discussions, as exemplified by the World Health Organization active engagement in COP26^1^ and the extraordinary joint publication across 200 health journals of a call for urgent climate action to protect health.^2^ To mitigate the effects of climate change for current and future generations, countries need to cut their emissions of greenhouse gas (GHG) and reach the global target of net zero emissions by the middle of the 21st century.^3^ These efforts could potentially result in important short-term health benefits, especially by improving air quality, housing, and diets, and increasing physical activity.^4^ Demonstrating health co-benefits of climate mitigation actions through a rigorous assessment could help make the case for transformative strategies and play a key role in increasing adhesion and commitment of people and their governments in reducing GHG emissions.^5,6^

Government and civil societies can contribute important efforts in exploring and understanding desirable pathways toward carbon neutrality. In France, governmental agencies, non-profit organizations, and other groups of qualified experts developed several carbon neutrality scenarios. These transition scenarios describe various combination of changes in socio-economic systems that are compatible with achieving a climate target. Based on a transdisciplinary approach, these scenarios provide explicit details on the scope and distribution of reduction efforts and on the role of the different levers of change, whether political technological or behavioural.^7,8^

In Europe, transport represents the second sector contributing the most to GHG emissions.^9^ In France in particular, transportation contributes to 31% of the national GHG emissions, making it the highest emitter. With about half of these emissions attributable to individual motorized vehicles,^10^ modal shifts towards low-carbon transport modes such as public or active transportation constitute an important element for national decarbonisation pathways.

There is compelling evidence that sedentary lifestyles cause a large health burden, especially in high-income countries, and that increasing physical activity could provide important public health benefits ^11,12^. Specifically, the potential for active transport modes such as cycling and biking to alleviate mortality and morbidity are now well established.^13,14^ Therefore, climate change mitigation actions resulting in increasing active transportation may carry important public health benefits.^4,15,16^ However, these benefits have not been quantified, yet, within the framework of a transition scenario describing detailed and credible societal transformations toward carbon neutrality.

In this study, we aimed at quantifying the health benefits of physical activity caused by active transportation on all-cause mortality in a French transition scenario over the 2021-2050 period. To do so, we relied on the framework of health impact assessment and follows recent guidelines for modelling and reporting health effects of climate change mitigation actions.^17,18^

## METHODS

### The negaWatt scenario

Since 2003, the negaWatt association develops energy transition scenarios for metropolitan France. These scenarios offer coherence in energy flows and a high level of sectoral detail while providing explicit data on the main physical determinants of energy consumption (heated surfaces, km-passengers) with a yearly time resolution. The latest negaWatt scenario was released in 2021.

In order to reach net zero emissions in 2050, the negaWatt approach combines energy sufficiency (favouring behaviours and activities that are intrinsically low on energy use, at individual and collective levels),^19^ energy efficiency and renewable sources. For the transportation sector, the scenario classifies trips based on their motives (commuting, occasional leisure, professional, etc.), lengths, and locations (urban centres, rural areas, etc.).

The negaWatt scenario activates several levers to decarbonize the transportation sector.^20^ First, the scenario assumes an overall decrease in transportation demand, mostly driven by a decrease in long-distance trips, an increase in teleworking, and an overall relocation of activities. Then, ambitious fiscal measures would integrate all externalities of transportation (CO2, air pollution, noise) while accounting for social and territorial inequalities issues. These measures would allow important investments in public and active transportation infrastructures, such as railways and bike lanes. These would yields to large modal shifts from individual motorized vehicles toward public and active transportation, especially for short trips. Motorized vehicles would remain the most common transport mode in terms of km-passengers (Supplementary figure 1), but decarbonisation is achieved assuming a reduction of cars weights, a higher vehicle occupancy rate (carsharing), and lastly a shift toward renewable energy, such as electricity and biogas.

The evolution of walking and cycling mileage between projected in the negaWatt scenario for 2021-2050 is presented in Figure 1. The weekly number of kilometres walked per inhabitant increases slightly and regularly, with an overall increase of +11% over the study period. The distance cycled increases most dramatically between 2021 and 2040, from 2.4 km.inh^-1^.week^-1^ to 17.1 km.inh^-1^.week^-1^ (+612%), it peaks at 17.5 km.inh^-1^.w^-1^ in 2045, and then decreases back to 17.1 km.inh^-1^.week^-1^ in 2050. This global increase of distances cycled is distributed between classical bike and E-bike, with an overall contribution of E-bikes increasing from 3.3% of the kilometres cycled in 2021 to 70% in 2040, this later proportion being kept constant over 2040-2050. The increase in distances cycled together with the relative decrease in the contribution of classical bikes explains the inverse U-shape peaking in 2032 that is observed for classical bike. As a comparison, the average distance travelled by car (regardless of the energy used) decreases by nearly 40% between 2021 to 2050 (from approximatively 12,000 to 7,500 km.inh^-1^.y^-1^).

**Figure 1:**
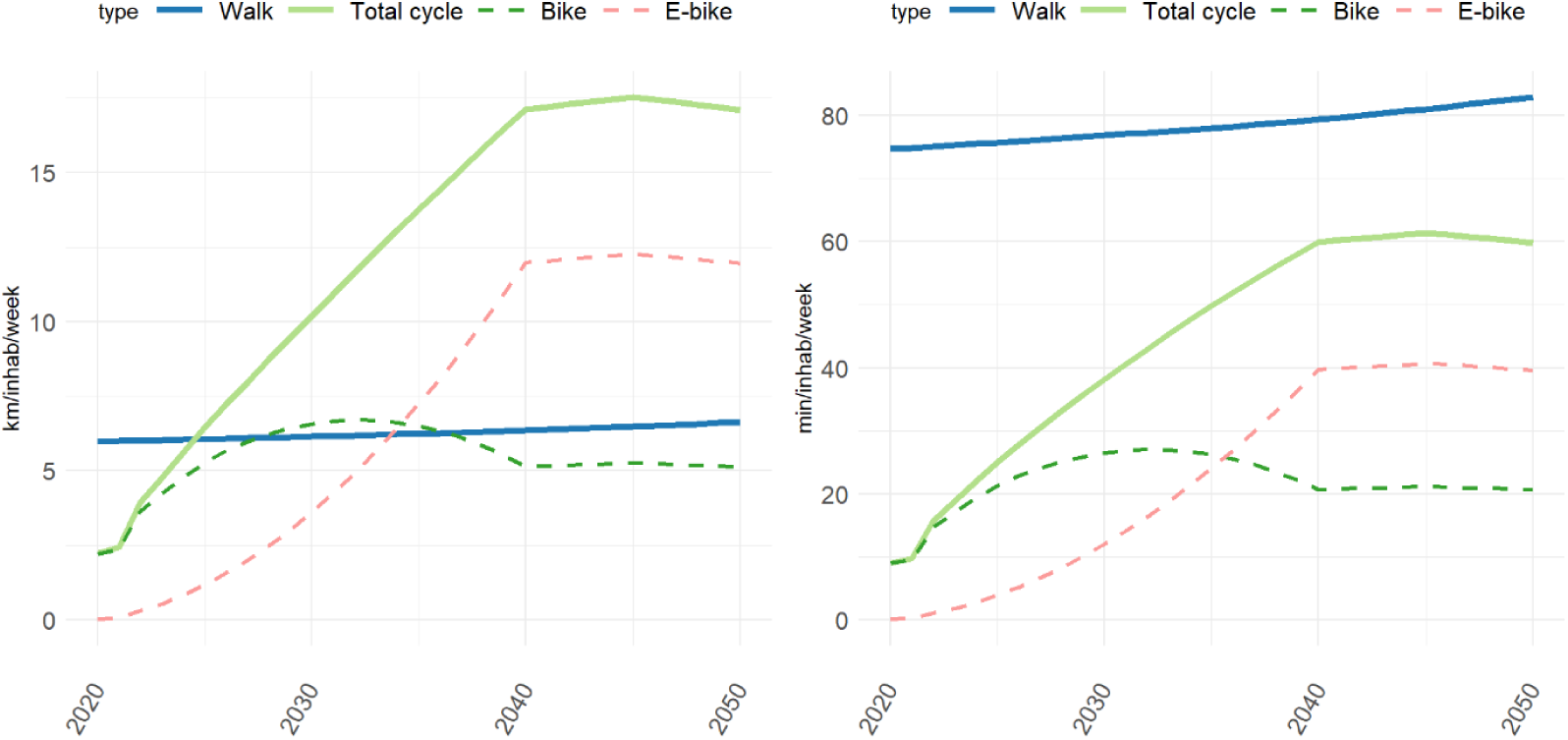
Evolution of weekly walking and cycling mileage (left) and duration (right) (negaWatt scenario, 2020-2050). Calculation of duration assumed an average speed of 4.8 km.h^-1^, 14.9 km.h^-1^, 18.1 km.h^-1^ for walking, cycling and E-biking, respectively.

### Age-distribution of cycling

There is currently a scarcity of reliable data regarding the frequency, volume (mileage) and age-distribution of active transportation in France. Results of the most recent nationally representative surveys have been released in early 2022, but at the time we conducted this analysis, the most recent nationally representative data was collected in 2008 and documented a biking mileage that was largely concentrated in young men aged <25 years.^21^ Investment in active transportation promotion and infrastructure, as well as technological development (notably E-bikes) are expected to have flattened this distribution. Moreover, a large increase in active transportation such as the one projected in the negaWatt scenario implies that all age groups would be affected. We thus used the age-distribution of walking and cycling mileage reported from Denmark, one of the country with the highest cycling level in Europe, as a target distribution to allocate the yearly global mileage projected by the negaWatt scenario across ages. The Danish age-distribution of active transportations mileage for 2016-2019 was obtained from the Danish National Travel Survey.^22^ We assumed the overall relative increase in walking and cycling to affect all ages homogeneously. However, for cycling, we attributed the total number of kilometres cycled between E-bike and classical bike differentially across age group in order to both reproduce : i) the 6.7 years difference observed in the mean age of E-bikers and cyclists reported in a recent multi-country study,^23^ and ii) the time-changing relative contribution of E-bike to the total cycling mileage assumed in the negaWatt scenario. Further details and formulas are detailed in Supplementary Text 1. For the reference (or business as usual) scenario we assumed the same relative age-distribution of active travel as detailed above, but the total mileage of active travel was assumed to be the same as in our base year, 2021.

### Health Impact Assessment

The scope of the study is metropolitan France and the time-period of the analysis is 2021-2050. Projection of demographic data for population size, age distribution and mortality rates used in the negaWatt scenario and in the HIA were obtained from the National Institute of Statistics and Economic Studies.^24^

We relied on the framework of the Health Economic Assessment Tool (HEAT) developed by the World Health Organization to estimate the number of deaths prevented by active transportation.^25^ Based on the meta-analysis by Kelly et al for the reduction in all-cause mortality from walking and cycling,^14^ we assumed a reduction by 10% in all-cause mortality (95% confidence interval, CI: 6-13%) for a weekly exposure of 100 minutes of cycling, and a reduction of 11% (95% CI: 4-17%) for a weekly exposure of 168 minutes of walking. Both of these reference volumes represent a metabolic equivalent of task hours per week of 11.25 MET.hour.week^-1^, independently of other physical activity, which correspond to the level of physical activity recommended by international guidelines.^26^ As the meta-analysis by Kelly et al considered all-cause mortality, negative sides of active transportation via road safety and increased exposure to air pollution were controlled for. We assumed that the physical activity required for an E-bike was 90% of the one required for a classical bike.^16^ Risk reduction estimates were scaled at the yearly level of the corresponding active travel using a linear dose-response function (DRF) which was capped at a maximum level of risk reduction (45% for cycling and 30% for walking).^14^ In the main analysis, the age range considered for a reduction of mortality attributable to physical activity increase was 20-84 years. Younger and older ages were disregarded because the evidence for the health effects of physical activity was not as large as that for the 20-84 years.

Yearly distances travelled by cycling, E-bike or walking were converted into times of exposure considering an average speed of 14.9 km.h^-1^, 18.1 km.h^-1^ and 4.8 km.h^-1^, respectively.^16,25^ We assumed the same values for E-bike speed and E-bike-to-classical-bike MET ratio as those recently identified in the literature by Egiguren et al.^16^ The scaled values of risk reduction were then applied to the projections of the age-specific (1-year age bands) mortality rate in both (ie. negaWatt and reference) scenarios for each year from 2022 to 2050. This allowed to estimate, for both scenarios, the yearly number of premature deaths averted by active transportation for each age. Based on the yearly projected value of life-expectancy, we also calculated the years of life lost (YLL) averted by active transportation associated with each scenario and for each travel mode, age and year. The health impact of the increase in active transportation in the negaWatt scenario as compared to the reference scenario was estimated as the difference of the number of deaths averted and YLL between the negaWatt and the reference scenario. We lastly used age- and year-specific death rates in each scenario to estimate the increase in life expectancy due to active transportation in the negaWatt scenario.

### Health economics evaluation

To estimate the economic health benefits of active transportation implied by the negaWatt scenario, we used the standard value of statistical life year (VSLY) that was recommended in 2013 in France for the socioeconomic evaluation of public investments.^27^ The recommended VSLY was 139k€ in 2020, and the values recommended for projections account for economic growth (publicly available here: https://www.strategie.gouv.fr/sites/strategie.gouv.fr/files/atoms/files/20181214_complement_b_valeurs_tutelaires.xlsx). All amounts were expressed in Euros 2020.

### Sensitivity analysis

We conducted several alternative analyses in order to assess the sensitivity of our results to specific parameters values or assumptions. The various alternative assessments we conducted considered : i) no age difference between classical bike and E-bike users; ii) a ratio of MET.hour of 0.78 for the physical activity required for an E-bike vs a classical bike, as assumed by Bouscasse et al;^28^ iii) an age limit of 75 years above which physical activity doesn’t decrease the mortality risk ; iv) a reduction by 19% (95% CI: 9% - 29%) of the risk of mortality for an exposure of 11.25 MET.hour.week^-1^, as recently documented by the meta-analysis by Zhao et al,^29^ that we also used to scale the risk reduction for E-bike.

All estimates were provided together with uncertainty intervals (UI) calculated using the lower and upper bounds of the 95% CI reported for the reduction of the mortality risk for each active transportation mode. Analysis were conducted using the R software version 4.1.2. All codes are publicly available at: https://github.com/pbarban/HIA_Transport_Transition.

## RESULTS

The distribution of weekly active transportation durations across age groups is presented in Figure 2. Based on the shape of the distribution observed in Denmark, walking is distributed fairly homogeneously across age groups. The total cycling distribution is more heterogeneous, with the 20-29 years-old group contributing the most to the global mileage. Although the increase over time of walking and cycling is assumed to be proportional across each age group, the age-specific evolution is different when considering classical vs. E-bike. While E-bike exposure is projected to be low across all age groups in 2025, we assume the increases over time to be proportionally higher in the older vs. younger age groups in comparison with the increase in classic cycling. For instance, in 2045, 58% of distances cycled is attributed to E-bike for the 15-19 years-old group, and 81% among the 65-69 years-old group.

**Figure 2:**
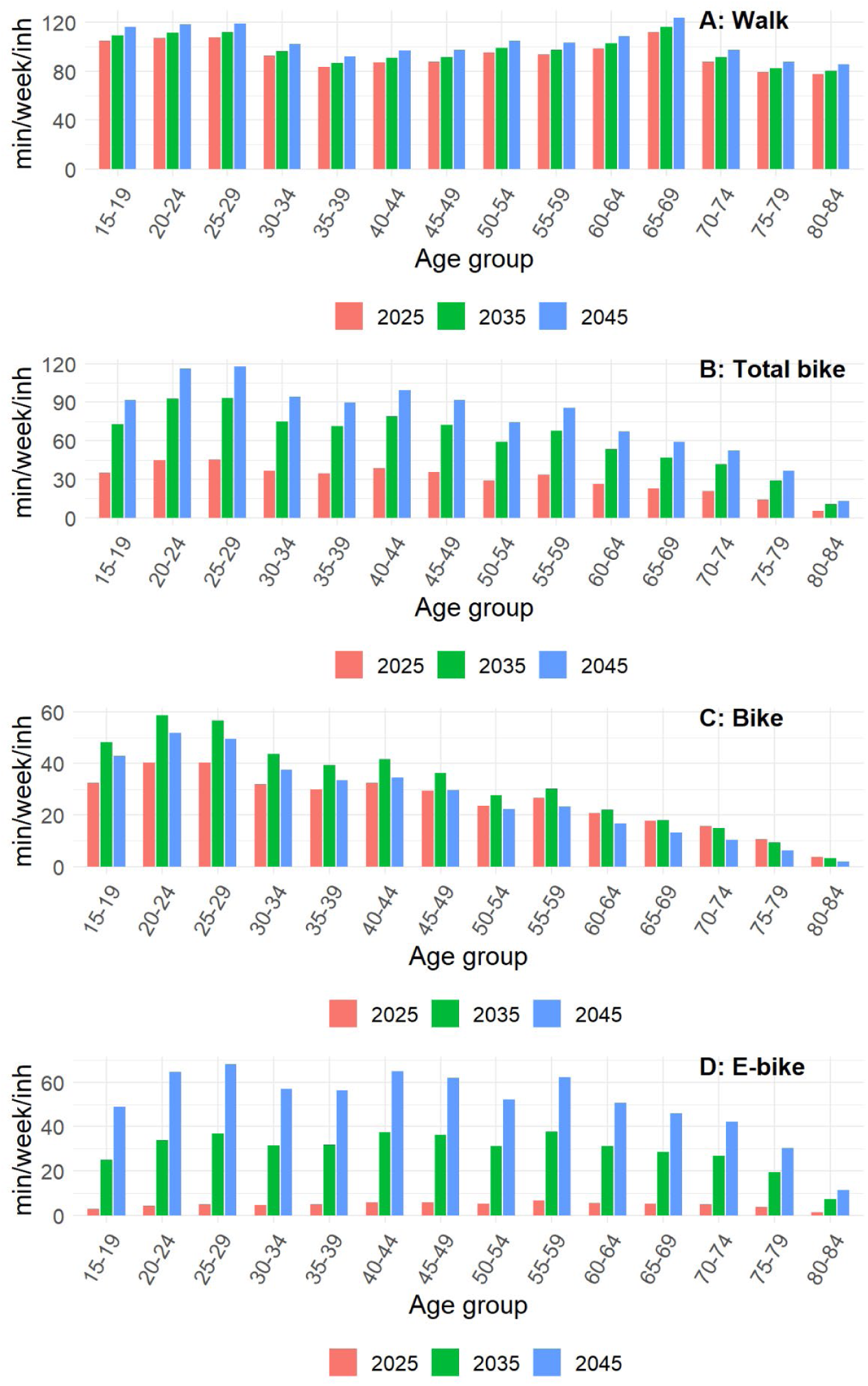
Evolution of weekly walking and cycling duration by age (negaWatt scenario, 2020-2050).

### Health impact assessment

The health impact estimated for active transportation over the 2021-2050 period are presented in Figure 3. As compared to a reference scenario in which walking and cycling mileage were maintained constant at their 2021 levels, the increased use of active transportation could prevent a cumulative 213,000 (UI: 122,000 -283,000) premature deaths across France from 2021 to 2050, with a yearly impact peaking at 9,825 [5,538-13,143] premature deaths prevented in 2045. The contribution of each transport mode varied across time (Figure 4). Over the whole study period, relatively to the cumulative number of death prevented, walking contributed to 11.3%, classical bike to 22.7%, and E-bike to 66.0%; these contribution were 8.7%, 27.9% and 63.4%, respectively, relatively to the cumulative number of YLL prevented.

**Figure 3:**
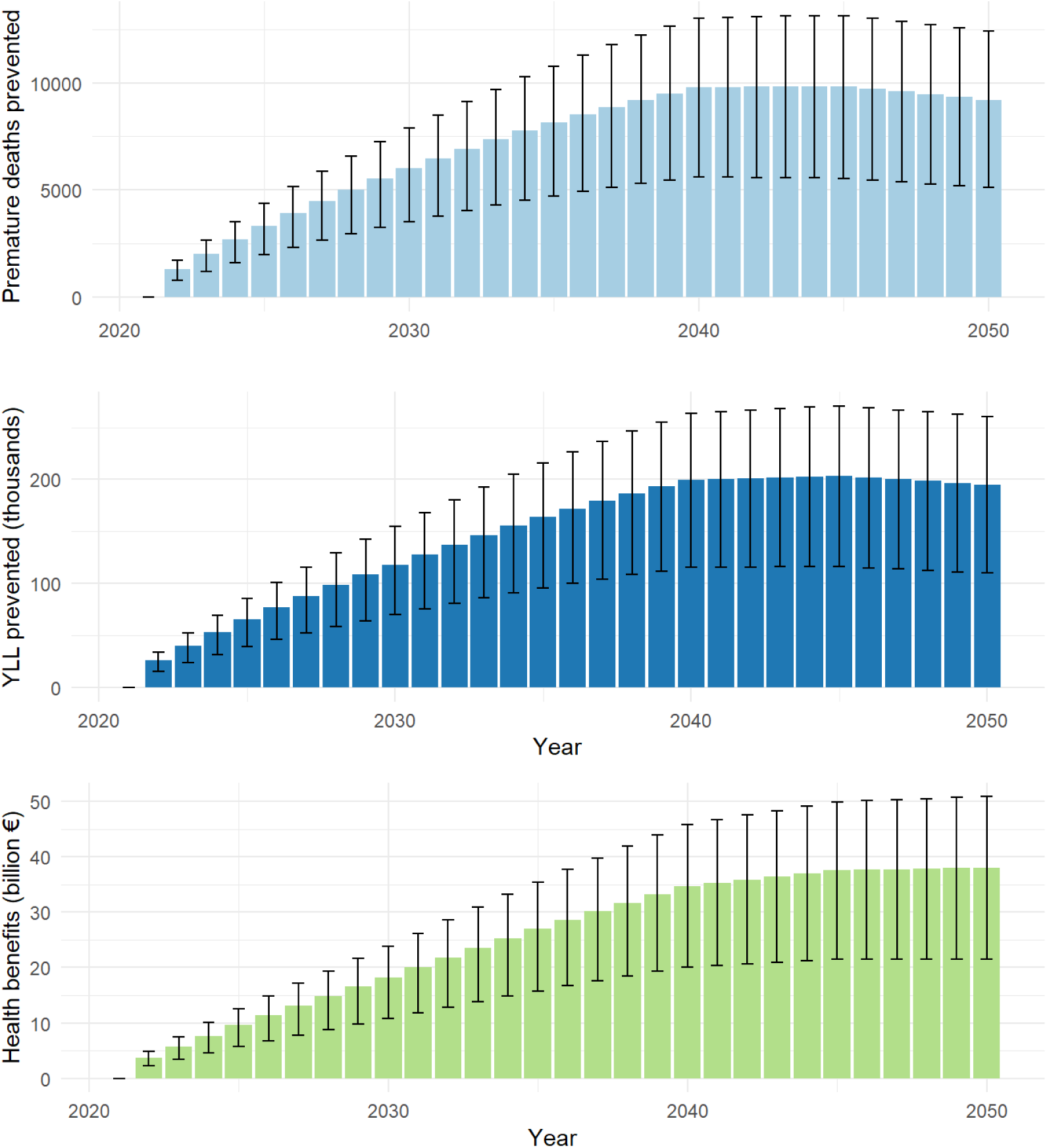
Premature deaths prevented (top), years of life lost (YLL) prevented (middle) and monetised health benefits (bottom) of active transportation (negaWatt scenario, 2020-2050). Health benefits are calculated based on the value of statistical life year.

**Figure 4:**
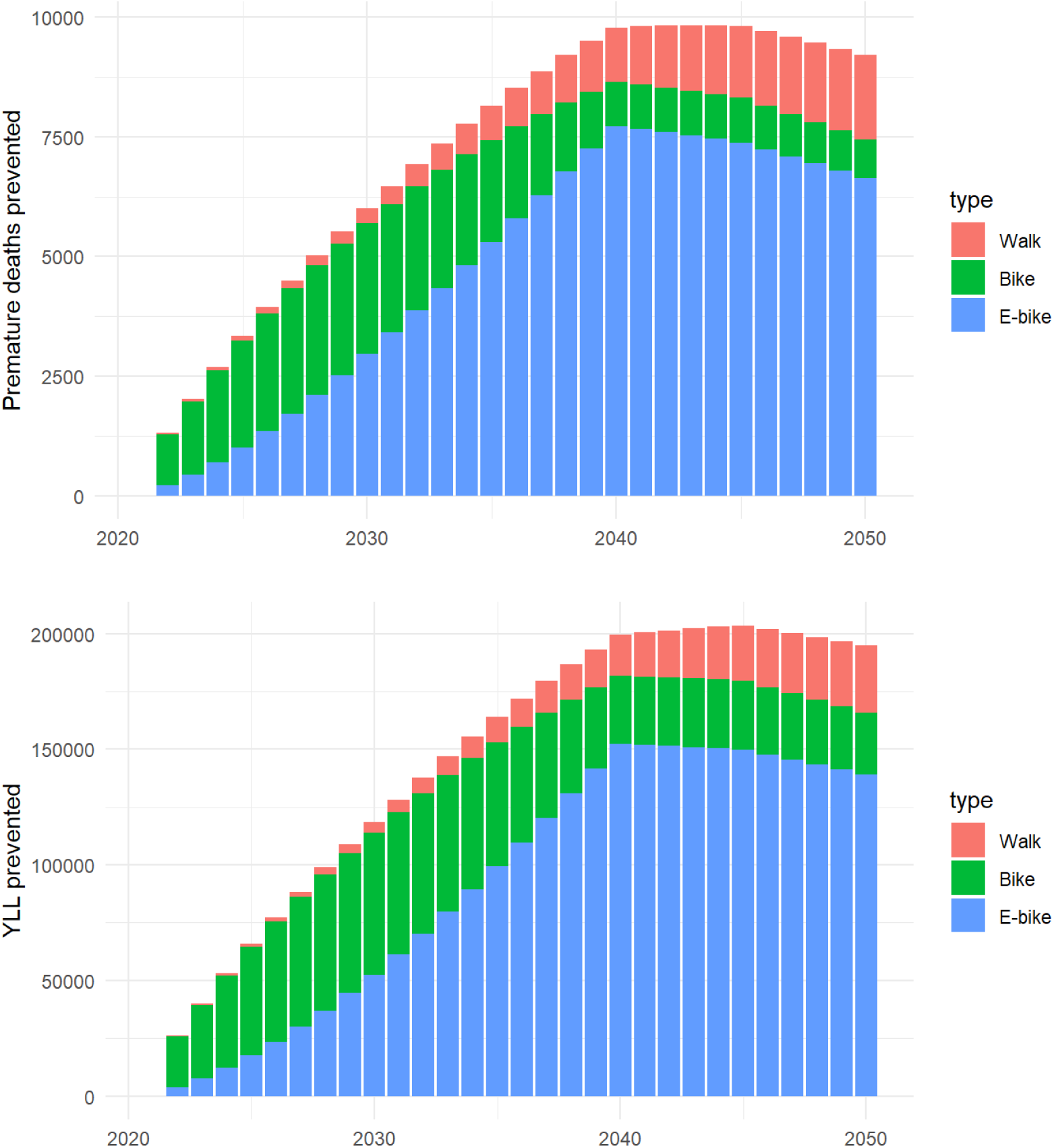
Premature deaths (top) and years of life lost (YLL, bottom) prevented per type of active transportation (negaWatt scenario, 2020-2050).

Death prevented were largely distributed among older age groups, especially among the 70-79 years-old group. For instance, in 2045, this age group concentrated 37.2% of premature death averted despite their contribution to only 9.1% of all km travelled, regardless of travel mode. In contrast, the distribution of YLL prevented was more widely distributed across age groups, and its mode was the 55-59 years-old group (Figure 5).

**Figure 5:**
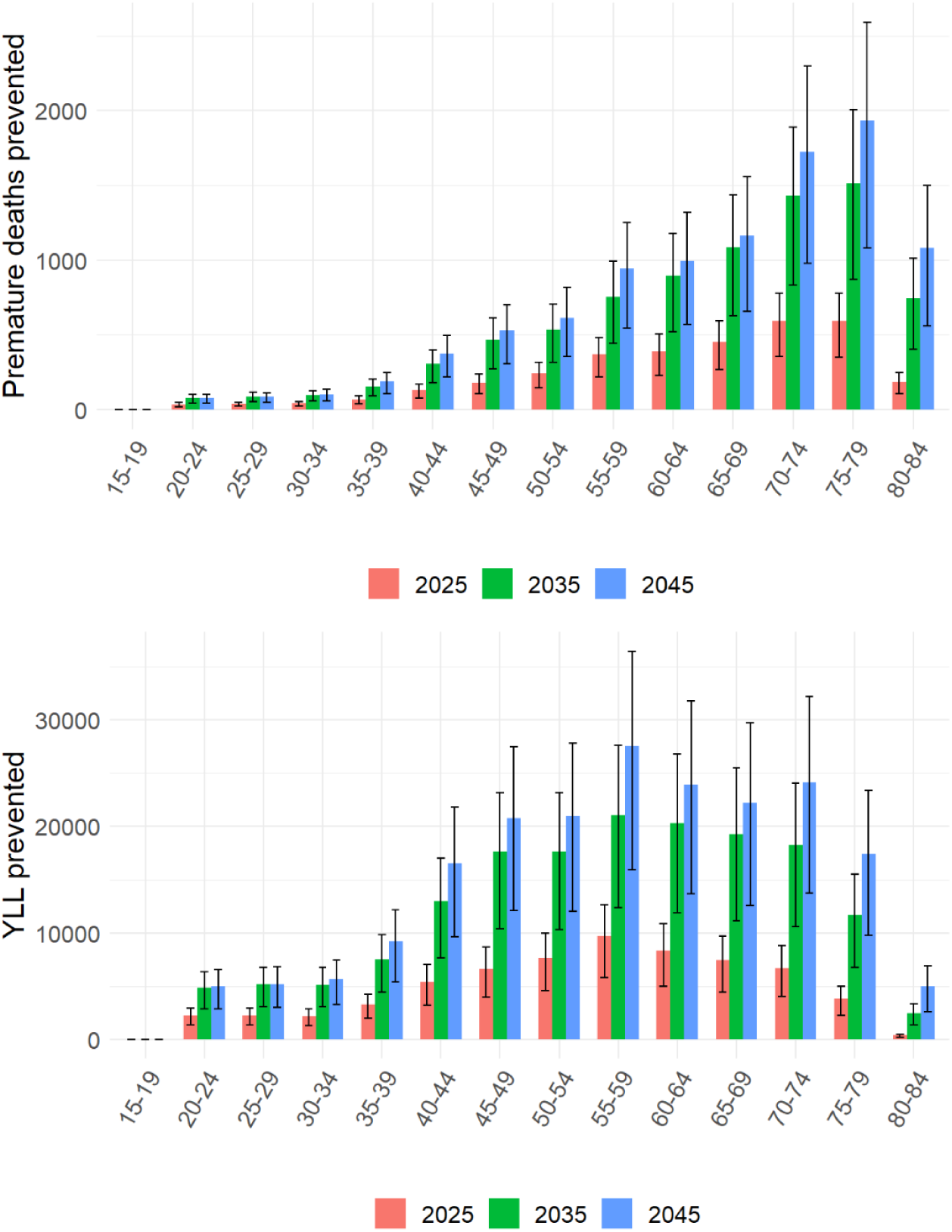
Contribution of the different age groups to premature deaths (top) and years of life lost (YLL, bottom) prevented by active transportation (negaWatt scenario, 2020-2050).

By considering age at death, we assessed that increased active transportation could prevent a cumulative 4.34 (UI: 2.52 – 5.74) millions of YLL over the study period), with a yearly amount peaking at 204 (UI: 116-271) thousands YLL prevented in 2045. We further estimated that the increased use in active transportation implied by the negaWatt scenario could result in a life expectancy gain of 3.39 (UI: 1.93-4.53) months over the general population in 2045 (Supplementary Figure 2).

Based on recommended values of statistical life year, we estimated the corresponding total economic health benefits that progressively increases to peak at €37.6 (UI: 21.5-49.9) billion in 2045, then plateauing after 2045. This represented a cumulative economic health benefits of €749 (UI: 433-990) billion over the 2021-2050 time period.

### Sensitivity analysis

Sensitivity of our results depending on the choice of the parameters are presented in Table 1. Assuming that there are no difference in age between bike vs E-bike users, or considering an alternative lower value of the E-bike-to-classical-bike MET ratio did not impact massively our results. Considering an age limit below 75 y. (vs. 85y. in the main analysis) to consider health benefits for physical activity decreased the estimated yearly and cumulative death prevented by approximatively 35%. However, the impact on the monetized health benefits was more limited (an approximated 10% decrease). Indeed, monetized health benefits calculation were based on YLL, and deaths prevented between 75 and 85 years are not those contributing the most to the overall YLL prevented by physical activity. Lastly, the parameters that appeared to have the largest impact on our results was the RR considered for bike (and consequently for E-bike as we maintained the same ratio for physical activity between bike and E-bike as in the main analysis). Using the more favourable value recently reported in the meta-analysis by Zhao et al^29^ leads to approximately doubling the estimates of health benefits, be it in terms of death prevented, YLL prevented, or life expectancy gain.

**Table 1:** Sensitivity analysis.

| Description | Premature deaths prevented |  | Years of life lost prevented |  | Monetised health benefits |  | Gain in life expectancy in 2045 |
| --- | --- | --- | --- | --- | --- | --- | --- |
|  | Annual, 2045 (UI) | Cumulative, 2021-2050, in thousands (UI) | Annual, 2045, in thousand (UI) | Cumulative, 2021-2050, in million (UI) | Annual, 2045, in billions € (UI) | Cumulative, 2021-2050, in billions € (UI) | In months (UI) |
| Main analysis | 9,825 [5538-13143] | 213 [122-283] | 204 [116-271] | 4.34 [2.52-5.74] | 37.6 [21.5-49.9] | 749 [433-990] | 3.39 [1.93-4.53] |
| No difference of mean age of classical vs. E-bike users | 10,144 [5730-13558] | 221 [127-293] | 208 [119-276] | 4.44 [2.57-5.86] | 38.3 [21.9-50.9] | 765 [443-1011] | 3.47 [1.98-4.63] |
| Ratio of E-bike MET to bike MET.hour = 0.78 (Bouscasse et al )<sup>28</sup> | 8,841 [4948-11864] | 195 [111-259] | 184 [104-245] | 3.98 [2.3-5.26] | 33.9 [19.3-45.1] | 684 [395-906] | 3.06 [1.73-4.09] |
| No protective health effects of physical activity above 74 y old | 6,402 [3667-8503] | 143 [83-189] | 176 [101-234] | 3.83 [2.23-5.05] | 32.5 [18.7-43.1] | 658 [382-869] | 2.75 [1.58-3.65] |
| Reported a value of 9% for 5MET.w-1, corresponding to RR=0.81 (Zhao et al)<sup>29</sup> | 17,409 [7846-26262] | 386 [175-582] | 367 [166-554] | 7.96 [3.62-11.99] | 67.8 [30.7-102.2] | 1,370 [622-2064] | 6.12 [2.75-9.26] |

RR : risk ratio, UI : uncertainty interval.

## DISCUSSION

In this study, we estimated the health impact of active transportation as projected in the negaWatt transition scenario, France. We estimate that increased use of walking and cycling could prevent up to 10,000 (between 5,000 and 13,000, approximatively) premature deaths and 190,000 YLL (between 105,000 and 250,000, approximatively) yearly in 2045. The uncertainty interval of these estimates reflects that surrounding the dose-response function linking physical activity and mortality. The health benefits could result in a life expectancy gain of about 3 months in 2045 for the general population. The health economic benefits generated by this burden alleviation could represent a cumulative €696 (between 403 and 919) billion over the 2022-2050 time period and an approximate €35 billion annually from 2045 onwards. To our knowledge, this study constitutes the first one to assess the health co-benefits of a credible pathway to net zero emission at the level of a country.

The negaWatt scenario we assessed assumes several mobility behavioural changes. First, it assumes a small but sustained increase of walking over the study period (+11%). This increase could be achieved by modal shifts from motorized vehicles to walking for short trips, or more broadly to public transportation, as this mode implies a certain amount of walking.^30^ The projected increase of cycling in the scenario is much larger (+612%). As France lags behind many countries on the current levels of cycling,^31^ the highest value of cycling volume projected for 2045 (62 min.week^-1^) in our scenario was still not dramatically higher than current average cycling in Denmark (45 min.week^-1^) and remains below the average cycling duration in the Netherlands (74 min.week^-1^ in 2010-2013).^22,32^ Moreover, this overall increase in cycling was mostly driven by E-bikes, projected to represent up to 70% of all kilometres cycled from 2040 onwards. Indeed, according to recent studies E-bike constitutes a promising transportation mode for many, especially for older adults, allowing longer trips than classical bikes and thus having a fair potential for car trips substitution.^23,33,34^

Cycling, either classical or E-bike, represented the largest health benefits in our study. Our results were therefore sensitive to the DRF chosen to quantify reductions of all-cause mortality by increased cycling. Our main analysis, relied on a DRF reported in a meta-analysis synthetizing the results 7 prospective studies and widely used in cycling-related HIAs.^25,28,32,35^ The more favourable DRF documented by the recent meta-analysis by Zhao et al used in our sensitivity analysis led to a doubling of the health impact estimates. Our approach built on the HEAT framework by explicitly accounting for age.^25^ In a conservative assumption, HEAT assumes no reduction in mortality due to physical activity beyond 65 years, while our main analysis assumed such a preventive effect up to 84 years. Indeed, both meta-analyses used in our assessment included studies providing data beyond age 65. Moreover, there is increasing evidence that regular physical activity in older people can reduce the risk of falls and fractures, and thus may reduce mortality.^36^

Several studies have previously assessed the health benefits of increased active transportation for France. Praznoczy estimated in 2012 that physical activity induced by a shift of 20% of modal share toward biking could prevent ∼3,000 premature deaths in 2020 in Ile-de-France (12.2 million inh., ∼20% of the population considered here).^37^ More recently, Egiguren et al estimated that a high-biking scenario could prevent ∼2,600 annual premature deaths in France in 2030.^16^ However, direct comparisons with our results are not straightforward as these studies relied on different geographical scope and on various assumptions, especially regarding the evolution of cycling mileage and modal share. In addition to the existing literature, our study highlights the potential of active transportation promotion policies to yield important public health benefits in the medium and long term.^38^ We estimated that active transportation implied by the negaWatt scenario may prevent up to 10,000 deaths annually from 2045 onwards. For comparison, efforts in road safety made in France over the past 10 years may have prevented about 1,500 deaths yearly ; and public health policies aiming at decreasing alcohol consumption by 20% in France would result in approximatively 7,000 deaths prevented a year.^39,40^

This assessment carries various limitations. On the one hand, we estimated the health impact of the negaWatt scenario as compared to a business-as-usual scenario that considered the level of active transportation constant from 2021 onward. Disregarding possible increases in cycling, as those having been reported in many French cities may have conducted to overestimated health impact.^41^ However, these recent increases have been partly linked to conjectural factors, such as the Covid-19 impact on public transportation, and one can question there durability in the absence of dedicated policies. On the other hand, the negaWatt scenario assumes a strong and continuous increase of cycling between 2021 and 2040. This increase could be partly hampered by increasing frequency and duration of heat waves that are predicted in the near and medium term due to climate warming.^42^ Our study assessed the health impact of active transportation for those exposed to these modes only. Pedestrian and bike-users are also exposed to negative health outcomes, such as increased risk of traffic injury and increased exposure to air pollution. The DRF for all-cause mortality we used captures both positive and negative effects of cycling and walking. The present analysis therefore implicitly accounted for the detrimental effects of walking and cycling. However, active transportation also carries altruistic health benefits beyond the groups of pedestrians and bike-users, such as reduced overall air or noise pollution. These altruistic benefits are not included in the present analysis, mostly for methodological issues. The health benefits we document here may thus have been underestimated. Indeed, In a multi-country analysis, Hamilton et al estimated that the benefits resulting from improve air quality implied by the Paris Agreement may be of the same order of magnitude than those resulting from active travel (although this ratio varied largely across countries).^4^

This study documents that physical activity due to increased active transportation projected in a transition scenario would generate substantial public health benefits, which may be comparable to the gains expected by large-scale health prevention interventions. The present study shows that these health gains may translate into important monetised health benefits on the short term, thus providing further arguments in favour on ambitious climate mitigation investment and policies. The fact that an energy transition scenario assuming increasing active transportation may generate health benefits is itself unsurprising and expectable. However, the rigorous quantification of such benefits offers more detailed elements, especially to weights the costs of policies promoting active transportation with regards to their benefits. Furthermore, several levers may be activated to decarbonize the transportation sector. For instance, some other French transition scenarios rely much more widely on the electrification of the vehicle fleet than the negaWatt scenario does.^43^ The present study thus suggests than betting on vehicle electrification only may miss the opportunity of large public health benefits generated by increased physical activity.

The primary novelty of this study is that our estimates are based on detailed and internally consistent scenario detailing an explicit pathway toward carbon neutrality at the scale of a country. The negaWatt association has developed these energy scenarios since 2003 and their influence on government policies have grown since then. Government agencies such as the French Agency for ecological transition have now produced energy scenarios quite similar to the negaWatt scenarios, in particular with regards to growth in active transportation. Previous similar health impact assessment of ambitious climate targets consisted in evaluating co-benefits of these targets once they were reached, with no consideration of the pathway.^4,15^ This study is a demonstration that realistic scenarios to reach net zero for climate change purposes can generate benefits far beyond reductions in GHG only. This is the type of evidence that can help create alliances across sectors and garner support for much needed transitions.^6^ In other words, this contribution may help engage the climate change and public health scientific communities, along with the broader civic society, to promote health-enhancing pathways to net zero emissions.

## Data Availability

All data produced are available online at

https://github.com/pbarban/HIA_Transport_Transition

## Notes

### Competing Interest Statement

The authors have declared no competing interest.

### Funding Statement

This study did not receive any funding.

### Summary of Updates

Revised after peer-review

