## Supplementary Figure 1 for "Assessing the health benefits of physical activity due to active commuting in a French energy transition scenario"

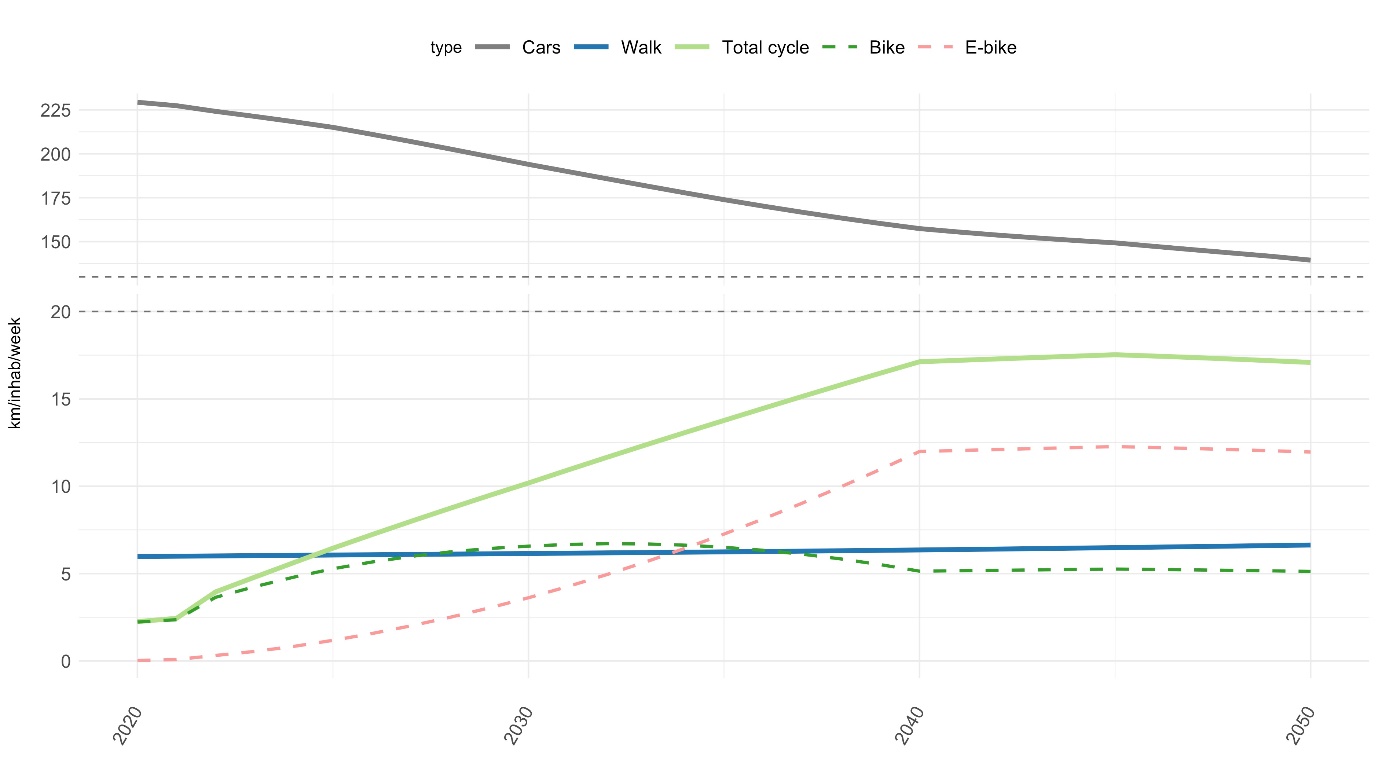

**Supplementary figure 1: Evolution of weekly mileage per transportation mode (negaWatt scenario, 2020-2050).**
