## Supplementary figure 2 for "Assessing the health benefits of physical activity due to active commuting in a French energy transition scenario"

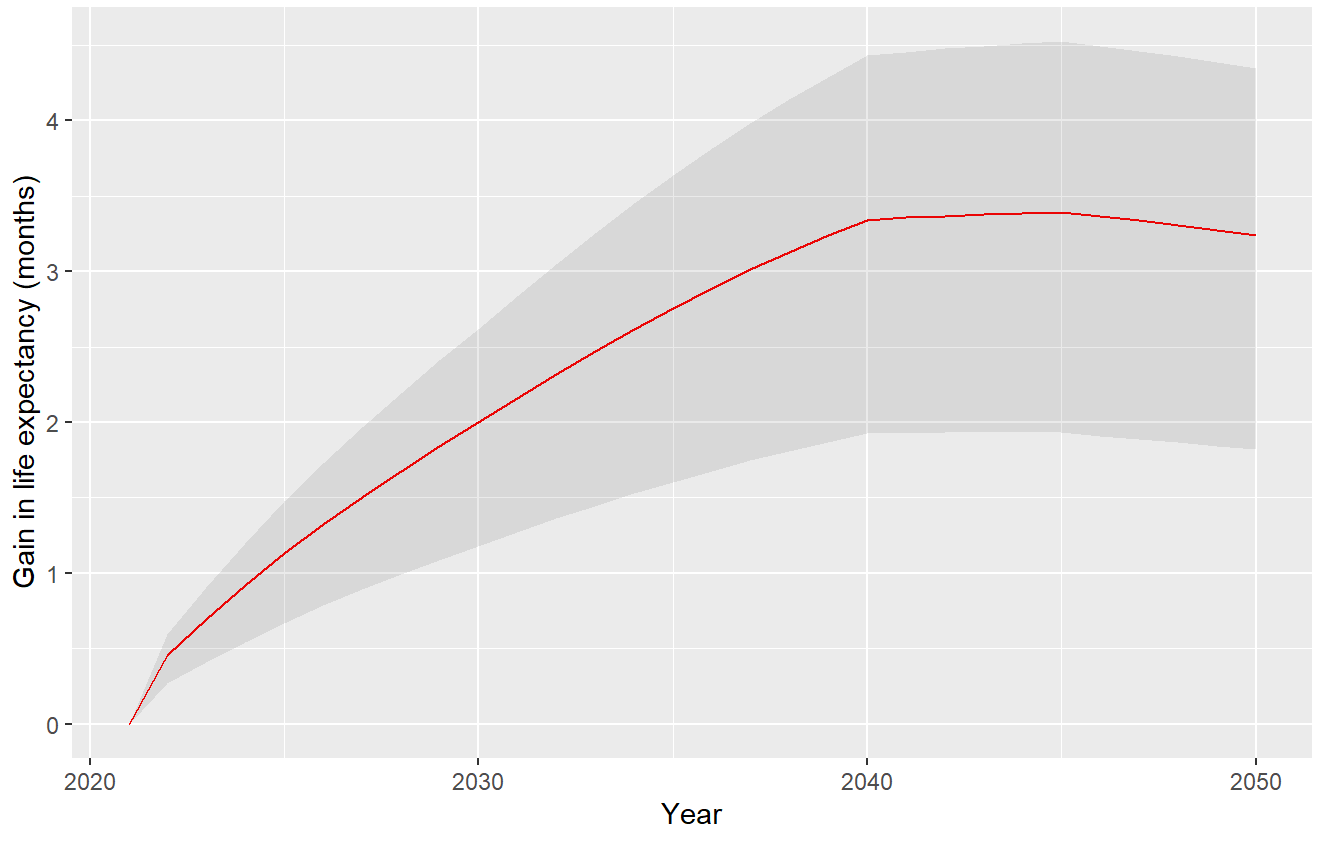

**Supplementary Figure 2 : Projected life expectancy gain, in months, expected by increased physical activity in the negaWatt scenario, 2020-2050 (grey: uncertainty interval).**
